# A decision aid for policymakers to estimate the impact of e-cigarette flavour restrictions on population smoking and e-cigarette use prevalence among youth versus smoking prevalence among adults

**DOI:** 10.1101/2022.11.14.22282288

**Authors:** Mark J Gibson, Marcus R Munafò, Angela S. Attwood, Martin J. Dockrell, Michelle A. Havill, Jasmine N Khouja

## Abstract

**Background:** Policy decisions should be evidence-based, but the magnitude of intended and unintended impacts cannot always be easily estimated from the available data. For example, banning flavours in electronic cigarettes (e-cigarettes) to reduce appeal to non-smoking young people could have the intended impact by reducing youth vaping but could have negative consequences for adult smokers and vapers.

**Methods:** We developed a decision aid to help policymakers make informed decisions on the potential net impact of a ban on e-cigarette flavours. We estimated the number of non-smoking youth who would be deterred from ever vaping and subsequently ever smoking, and the number of smokers and ex-smokers who would be deterred from quitting or encouraged to relapse, to determine whether the benefits to youth outweigh the costs to existing smokers and vapers. This aid then outputs a report with the results graphically depicted to aid interpretability.

**Results:** We demonstrated the value of this decision aid using data from various sources to estimate the impact of a flavour ban in three populations: the general UK population, low-socioeconomic position UK population, and the general US population. All three examples suggested a negative net population impact of a ban. These reports were then presented to the all-party parliamentary group for vaping.

**Conclusions:** We demonstrate how decision aids can be used to help policymakers arrive at evidence-based decisions efficiently and can be used to quickly obtain up-to-date estimates as new data becomes available.

---

Policy decisions should be evidence-based and lead to positive, beneficial impacts in the affected population. However, sourcing relevant evidence that can be easily interpreted can be a difficult task for policymakers working under time constraints. Creating decision aids for policymakers that can quickly provide brief, digestible guidance can be particularly useful in areas where existing evidence suggests the proposed policy change may have positive and negative implications on the target population.

One example is electronic cigarette (e-cigarette) policy. Some jurisdictions have banned flavours in e-cigarettes to reduce appeal to non-smoking young people and the UK could do the same; this could have the intended impact by reducing youth vaping but could have negative consequences for adult smokers and e-cigarette users (vapers). Although e-cigarettes are considered to be less harmful than cigarettes [1], and can be used by smokers to help them quit [2], there have been concerns that the wide range of available flavours encourage non-smoking youth to vape and subsequently smoke. While there is some evidence to suggest that flavours encourage youth vaping in both the US and the UK, there is no clear evidence that they encourage subsequent smoking [3-6]. The emergence of disposable vapes, which are most popular and relatively accessible among young people in both the US and UK, has further fuelled concerns about flavours in e-cigarette products [7, 8]. These concerns have led to bans of e-cigarette flavours (i.e., all but unflavoured, tobacco and menthol) in several jurisdictions. Evidence of the actual and predicted effect of bans is conflicting with some studies suggesting a reduction of vaping rates [9, 10] and others suggesting no reduction [11] or an increase in smoking rates in both youth [12] and adults [10, 13].

Contrasting evidence on the effectiveness of a potential ban makes it difficult for policymakers to reach an informed decision. Therefore, to help policymakers make informeddecisions on a potential e-cigarette flavour ban, we aimed to develop a decision aid for policymakers to specifically estimate the impact of a ban in any given population, and to illustrate the potential value of such decision aids in general.

## Decision Aid for Policymakers

We consulted with policymakers and researchers to create a decision aid to estimate the potential net impact of an e-cigarette flavour ban, for which population sizes and the proportions are used as inputs to estimate four numbers (See Figure 1 for calculations):

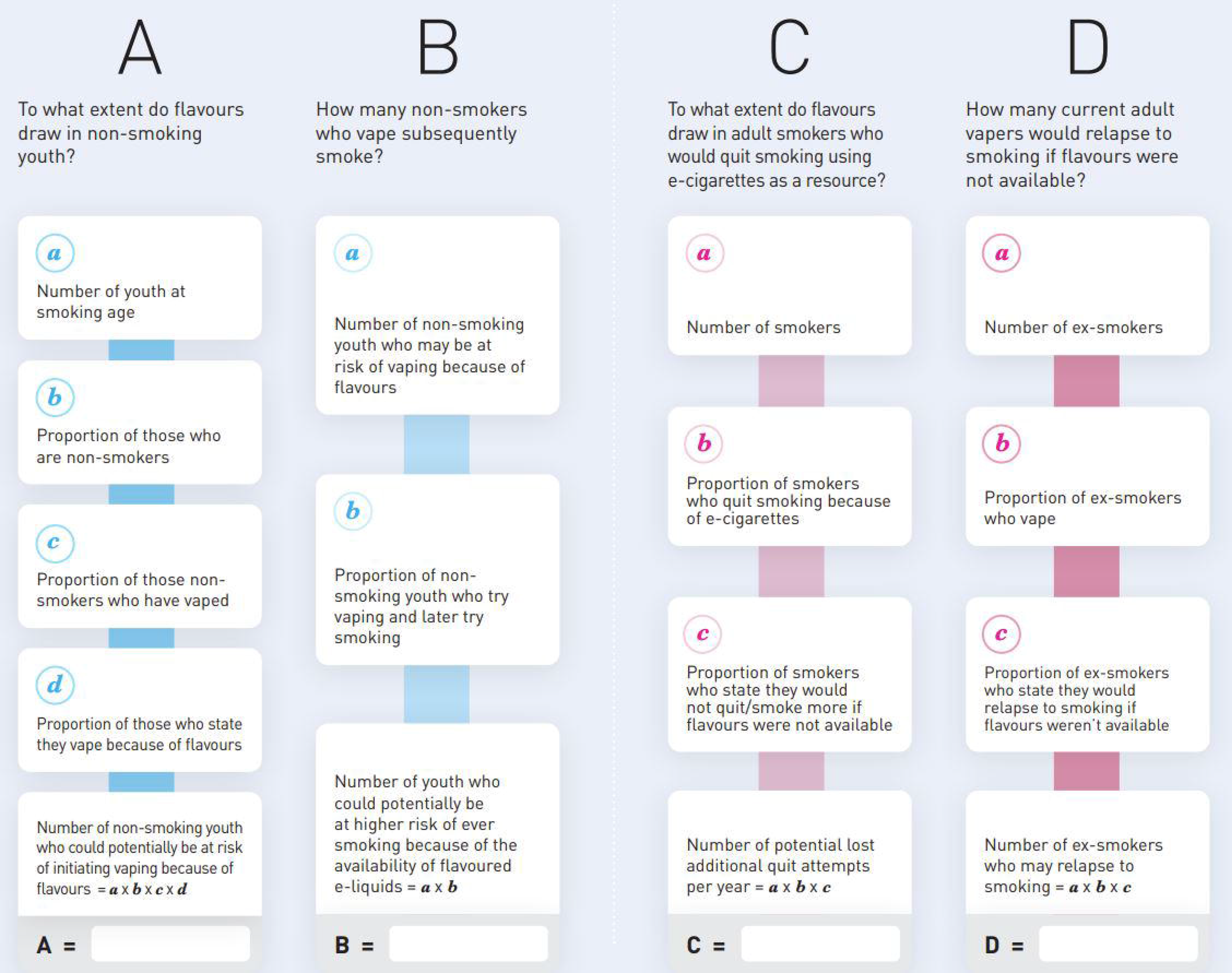

A. The number of non-smoking youth who could potentially be at risk of initiating vaping because of flavours.
B. The number of youth who could potentially be at higher risk of ever smoking because of the availability of flavoured e-liquids.
C. The number of potential lost additional quit attempts per year.
D. The number of ex-smokers who may relapse to smoking.

In this context, “youth” refers to those in the age range considered to be at risk of smoking (i.e.,11-17). From these four numbers the decision aid then calculates whether the number of non-smoking e-cigarette users introduced into the UK population because of flavoured e-liquid availability could outweigh the number of smokers and ex-smokers who might vape instead of smoke because of flavoured e-liquid availability (A – [C + D]) and whether the number of ever smokers introduced into the UK population because of flavoured e-liquid availability could outweigh the number of smokers and ex-smokers who might vape instead of smoke because of flavoured e-liquid availability (B – [C + D]). Positive values provide some support for a total ban on flavoured e-liquids, and negative values provide some evidence that the negative consequences of a total ban would outweigh the potential benefits (i.e., a negative net impact of a ban).

## Applied Examples

### Methods

We entered data from a variety of sources into our decision aid to estimate the effects of a smoking ban in the general UK population, the low-socioeconomic position UK population (specifically those from the C2, D and E categories of the National Readership Survey scale [14], which are those classified as skilled working class, working class or non-working) and the general US population. We used data from Action on Smoking and Health (ASH) [15], the Smoking Toolkit Study (STS) [16], the National Youth Tobacco Survey (NYST) [17] and publicly available figures from the Office of National Statistics (ONS) [18, 19], the Department for Work and Pensions [20], the Centres for Disease Control and Prevention (CDC) [21-23], census data [24-27], privately owned real-time statistics website Worldometer (estimates taken on 12/09/2022) [28, 29] and peer-reviewed academic research articles [30-32].

All population numbers or proportions and descriptions of data sources for these examples can be found in Supplementary Tables S1-3 and Supplementary Materials. Analysis code can be found at https://github.com/MRCIEU/Estimation-of-the-impact-of-flavour-restrictions-on-population-smoking-and-e-cigarette-use.

### Output

For the UK general population, we calculated that 53,609 non-smoking youth were at risk of ever vaping due to flavours and 26,269 of those were at risk of ever smoking due to flavours. 33,000 potential quit attempts would be lost a year and 295,403 ex-smokers would relapse to smoking.

For the UK low-socioeconomic position population, we calculated that 30,484 non-smoking youth were at risk of ever vaping due to flavours and 13,109 of those were at risk of ever smoking due to flavours. 19,096 potential quit attempts would be lost a year and 171,299 ex-smokers would relapse to smoking.

For the US general population, we calculated that 355,617 non-smoking youth were at risk of ever vaping due to flavours and 78,236 of those were at risk of ever smoking due to flavours. 172,481 potential quit attempts would be lost a year and 1,369,341 ex-smokers would relapse to smoking.

For all three populations, the decision aid estimated that the number of adult smokers and ex-smokers who choose not to quit or relapse would outweigh the number of youth vapers who would be deterred from vaping or smoking if a ban was introduced.

The final report produced by the decision aid for the general UK population (including the resulting guidance for policymakers) is shown in Supplementary Material.

## Discussion

We developed a decision aid to assist policymakers in decisions relating to a potential ban of e-cigarette flavours. We applied this decision aid to three different populations (with the output suggesting the ban would have a negative impact in the general UK, low-socioeconomic UK and general US populations). These reports were then presented to the UK All Party Parliamentary Group for Vaping (https://publications.parliament.uk/pa/cm/cmallparty/210602/vaping.htm), with the opportunity for clarification from, and feedback to, the researchers. This project demonstrates both the feasibility and utility of developing decision aids to increase the impact of science and facilitate the making of evidence-based decisions by policy makers.

While decision aids are useful, the output should always be used in combination with evidence from a range of studies and the interpretability of the output is dependent on the quality of the decision aid and the evidence supplied to it. There are some limitations of the decision aid demonstrated in this article. Firstly, much of the evidence is reliant on self-reported beliefs about how smokers and vapers think they will behave in the event of flavour restrictions being implemented, which may differ from their actual behaviour. Secondly, while there is a strong association between e-cigarette use and later smoking, there is no clear evidence that e-cigarette use *causes* smoking among youth. There is also evidence to suggest that at least part of this relationship could be explained by shared risk factors [33], so it is likely that this decision aid will overestimate the number of youth who are at risk of smoking due to e-liquid flavour availability. Furthermore, the decision aid does not account for displacement (i.e., the number of youths who do not smoke because e-cigarettes are available), meaning the benefit of a ban is likely overestimated [34, 35]. The decision aid also does not take into account negative consequences of a ban beyond smoking behaviour, such as the creation of a black market which could lead to the use of unregulated/homemade products [36] and death [37, 38]. It can also not predict the consequences of partial bans such as allowing flavoured e-liquid on prescription, banning only flavours which appeal to children or banning packaging which is appealing to children.

## Conclusions

The development of decision aids for policymakers can increase the impact of research and facilitate the making of evidence-based decisions by policymakers and the decision aid outlined in this article can inform on the net impact of an e-cigarette flavours ban. Using the currently available data, the decision aid demonstrated here suggests that a flavour ban would have a negative impact on the UK general, UK low socioeconomic and the US general populations. However, the output from the decision aid needs to be regularly updated to ensure it accurately represents the ever-changing sociocultural landscape. Future iterations should also investigate the effect of a potential ban on other at-risk populations (such as those who suffer from mental illness) and develop the decision aid to account for the effects of different e-cigarette flavours (i.e., sweet versus not), and the effect of disposables, as the relevant data becomes available.

## Data Availability

All data sources are outlined in the supplementary tables (with links) and are public or available upon request.

## Declarations

### Ethical approval and consent to participate

This study only used secondary data. All methods were carried out by the original studies in accordance with relevant guidelines and regulations and informed consent was obtained from all subjects and/or their legal guardian(s). Ethical approval for the Smoking Toolkit Study was granted by the UCL ethics committee (ID 0498/001). All other data is publicly available or available upon request.

### Consent for publication

NA

### Availability of data and materials

The Smoking Toolkit Study data can be requested at https://smokinginengland.info/resources/sts-documents. Action on Smoking and Health can be contacted for data requests at https://ash.org.uk/contact-us. The National Youth Tobacco Survey data can be downloaded at https://www.cdc.gov/tobacco/data_statistics/surveys/nyts/data/index.html

### Author contributions

The policy decision aid was designed by MRM, ASA, MJD, MAH and JNK who also provided feedback on this manuscript; The applied examples were conducted and the manuscript was written by MJG under the supervision of JNK.

## Acknowledgments

We would like to thank the Smoking Toolkit Study and Smoking on Action Health for providing data.

## References

1. McNeill A, Brose LS, Calder R, Hitchman SC, Hajek P, McRobbie H. E-cigarettes: an evidence update a report commissioned by Public Health England. London: UK, PHE; 2015.

2. Hartmann-Boyce J, Begh R, Aveyard P. Electronic cigarettes for smoking cessation. Bmj-Brit Med J. 2018;360.

3. Pepper JK, Ribisl KM, Brewer NT. Adolescents’ interest in trying flavoured e-cigarettes. Tob Control. 2016;25:ii62–ii6.

4. Notley C, Gentry S, Cox S, Dockrell M, Havill M, Attwood AS, et al. Youth use of e-liquid flavours-a systematic review exploring patterns of use of e-liquid flavours and associations with continued vaping, tobacco smoking uptake or cessation. Addiction. 2022;117(5):1258–72.

5. Chan GCK, Stjepanovic D, Lim C, Sun TZ, Anandan AS, Connor JP, et al. Gateway or common liability? A systematic review and meta-analysis of studies of adolescent e-cigarette use and future smoking initiation. Addiction. 2021;116(4):743–56.

6. Khouja JN, Suddell SF, Peters SE, Taylor AE, Munafo MR. Is e-cigarette use in non-smoking young adults associated with later smoking? A systematic review and meta-analysis. Tob Control. 2021;30(1):8–15.

7. Park-Lee E, Ren CF, Sawdey MD, Gentzke AS, Cornelius M, Jamal A, et al. E-Cigarette Use Among Middle and High School Students - National Youth Tobacco Survey, United States, 2021. Mmwr-Morbid Mortal W. 2021;70(39):1387–9.

8. Tattan-Birch H, Jackson SE, Kock L, Dockrell M, Brown J. Rapid growth in disposable e-cigarette vaping among young adults in Great Britain from 2021 to 2022: a repeat cross-sectional survey. Addiction. 2022;Accepted Articles.

9. Rogers T, Brown EM, Siegel-Reamer L, Rahman B, Feld AL, Patel M, et al. A Comprehensive Qualitative Review of Studies Evaluating the Impact of Local US Laws Restricting the Sale of Flavored and Menthol Tobacco Products. Nicotine & Tobacco Research. 2022;24(4):433–43.

10. Yang Y, Lindblom EN, Salloum RG, Ward KD. The impact of a comprehensive tobacco product flavor ban in San Francisco among young adults. Addictive Behaviors Reports. 2020;11:100333.

11. Romm KF, Henriksen L, Huang JD, L. D, Clausen M, Duan ZS, et al. Impact of existing and potential e-cigarette flavor restrictions on e-cigarette use among young adult e-cigarette users in 6 US metropolitan areas. Preventive Medicine Reports. 2022;28.

12. Friedman AS. A Difference-in-Differences Analysis of Youth Smoking and a Ban on Sales of Flavored Tobacco Products in San Francisco, California. Jama Pediatr. 2021;175(8):863–5.

13. Khouja J, Dyer ML, Havill MA, Dockrell MJ, Munafo M, Attwood AS. Exploring the opinions and potential impact of unflavoured e-liquid on smoking cessation among UK smokers and smoking relapse among UK e-cigarette users: Findings from a qualitative study. Research Square. 2022.

14. National Readership Survey scale. Social Grade [Available from: https://www.nrs.co.uk/nrs-print/lifestyle-and-classification-data/social-grade/.

15. Sandford A. ASH. Trends in Urology & Men’s Health. 2012;3(2).

16. Fidler JA, Shahab L, West O, Jarvis MJ, McEwen A, Stapleton JA, et al. ‘The smoking toolkit study’: a national study of smoking and smoking cessation in England. Bmc Public Health. 2011;11.

17. Centers for Disease Control and Prevention. National Youth Tobacco Survey (NYTS) [updated 14/03/2022. Available from: https://www.cdc.gov/tobacco/data_statistics/surveys/nyts/index.htm.

18. Office of National Statistics. Smoking prevalence in the UK and the impact of data collection changes: 2020 2021 [updated 07/12/2021. Available from: https://www.ons.gov.uk/peoplepopulationandcommunity/healthandsocialcare/drugusealcoholandsmoking/bulletins/smokingprevalenceintheukandtheimpactofdatacollectionchanges/2020#.

19. Office of National Statistics. Smoking habits in the UK and its constituent countries 2021 [updated 07/12/2021. Available from: https://www.ons.gov.uk/peoplepopulationandcommunity/healthandsocialcare/healthandlifeexpectancies/datasets/smokinghabitsintheukanditsconstituentcountries.

20. Department for Work and Pensions. Households below average income: an analysis of the income distribution FYE 1995 to FYE 2020 [updated 25/03/2021. Available from: https://www.gov.uk/government/statistics/households-below-average-income-for-financial-years-ending-1995-to-2020/households-below-average-income-an-analysis-of-the-income-distribution-fye-1995-to-fye-2020.

21. Centers for Disease Control and Prevention. Current Cigarette Smoking Among Adults in the United States [updated 17/03/2022. Available from: https://www.cdc.gov/tobacco/data_statistics/fact_sheets/adult_data/cig_smoking/index.htm#.

22. Kramarow EA. Health of Former Cigarette Smokers Aged 65 and Over: United States, 2018. Centers for Disease Control and Prevention; 2020.

23. Centers for Disease Control and Prevention. Electronic Cigarette Use Among U.S. Adults, 2018 [updated 30/04/2020. Available from: https://www.cdc.gov/nchs/products/databriefs/db365.htm#.

24. United States Census Bereau. Quick Facts [updated 01/07/2021. Available from: https://www.census.gov/quickfacts/fact/table/US/PST045221.

25. Childstats.gov. POP1 CHILD POPULATION: NUMBER OF CHILDREN (IN MILLIONS) AGES 0–17 IN THE UNITED STATES BY AGE, 1950–2020 AND PROJECTED 2021–2050 [Available from: https://www.childstats.gov/americaschildren/tables/pop1.asp.

26. Statistica. Population of the United Kingdom from 1871 to 2020 [updated 05/09/2022. Available from: https://www.statista.com/statistics/281296/uk-population/.

27. Statistica. Population of the United Kingdom in 2020, by age [updated 05/09/2022. Available from: https://www.statista.com/statistics/281174/uk-population-by-age/.

28. Worldometer. United States Population [updated 12/09/2022. Available from: https://www.worldometers.info/world-population/us-population/.

29. Worldometer. U.K. Population [updated 12/09/2022. Available from: https://www.worldometers.info/world-population/uk-population/.

30. Berry KM, Fetterman JL, Benjamin EJ, Bhatnagar A, Barrington-Trimis JL, Leventhal AM, et al. Association of Electronic Cigarette Use With Subsequent Initiation of Tobacco Cigarettes in US Youths. Jama Network Open. 2019;2(2).

31. Chen RF, Pierce JP, Leas EC, Benmarhnia T, Strong DR, White MM, et al. Effectiveness of e-cigarettes as aids for smoking cessation: evidence from the PATH Study cohort, 2017-2019. Tob Control. 2022.

32. Gravely S, Smith DM, Liber AC, Cummings KM, East KA, Hammond D, et al. Responses to potential nicotine vaping product flavor restrictions among regular vapers using non-tobacco flavors: Findings from the 2020 ITC Smoking and Vaping Survey in Canada, England and the United States. Addict Behav. 2022;125.

33. Khouja JN, Wootton RE, Taylor AE, Smith GD, Munafo MR. Association of genetic liability to smoking initiation with e-cigarette use in young adults: A cohort study. Plos Medicine. 2021;18(3).

34. Sokol NA, Feldman JM. High School Seniors Who Used E-Cigarettes May Have Otherwise Been Cigarette Smokers: Evidence From Monitoring the Future (United States, 2009-2018). Nicotine Tob Res. 2021;23(11):1958–61.

35. Hallingberg B, Maynard OM, Bauld L, Brown R, Gray L, Lowthian E, et al. Have e-cigarettes renormalised or displaced youth smoking? Results of a segmented regression analysis of repeated cross sectional survey data in England, Scotland and Wales. Tob Control. 2020;29(2):207–16.

36. Action on Smoking Health. Use of e-cigarettes (vapes) among adults in Great Britain. 2020.

37. Werner AK, Koumans EH, Chatham-Stephens K, Salvatore PP, Armatas C, Byers P, et al. Hospitalizations and Deaths Associated with EVALI. New Engl J Med. 2020;382(17):1589–98.

38. Ellington S, Salvatore PP, Ko J, Danielson M, Kim L, Cyrus A, et al. Update: Product, Substance-Use, and Demographic Characteristics of Hospitalized Patients in a Nationwide Outbreak of E-cigarette, or Vaping, Product Use-Associated Lung Injury - United States, August 2019-January 2020. Mmwr-Morbid Mortal W. 2020;69(2):44–9.

