## Supplementary Materials for "A decision aid for policymakers to estimate the impact of e-cigarette flavour restrictions on population smoking and e-cigarette use prevalence among youth versus smoking prevalence among adults"

### E-LIQUID POLICY DECISION AID

University of BRISTOL

#### HEALTH PROTECTION OF FLAVOUR RESTRICTION

### A

To what extent do flavours draw in non-smoking youth?

**a** 6,409,450 of youth at smoking age

**b** 82% of those are non-smokers

**c** 6% of those non- smokers have vaped

**d** 17% of those state they vape because of flavours

Number of non-smoking youth who could potentially be at risk of initiating vaping because of flavours = a x b x c x d

**A =** 53,609

### B

How many non-smokers who vape subsequently smoke?

**a** 53,609 of non-smoking youth may be at risk of vaping because of flavours

**b** 49% of non- smoking youth who try vaping later try smoking

Number of youth who could potentially be at higher risk of ever smoking because of the availability of flavoured e-liquids = a x b

**B =** 26,269

#### HEALTH COST OF FLAVOUR RESTRICTIONS

### C

To what extent do flavours draw in adult smokers who would quit smoking using e-cigarettes as a resource?

**a** 5,500,000 smokers

**b** 2% of smokers quit smoking because of e-cigarettes

**c** 30% of smokers state they would not quit/smoke more if flavours were not available

Number of potential lost additional quit attempts per year = a x b x c

**C =** 33,000

### D

How many current adult vapers would relapse to smoking if flavours were not available?

**a** 17,479,451 ex-smokers

**b** 13% of ex-smokers vape

**c** 13% of ex-smokers state they would relapse to smoking if flavours weren't available

Number of ex-smokers who may relapse to smoking = a x b x c

**D =** 295,403

#### POLICY DECISIONS

### A-(C+D)

***Positive values*** indicate the number of youth vaping due to flavours outweighs the number of smokers and ex- smokers not smoking due to flavours.

***Negative values*** indicate the number of smokers and ex-smokers who are not smoking due to flavours outweighs the number of youth vaping due to flavours.

### B-(C+D)

***Positive values*** indicate the number of youth smoking due to flavours outweighs the number of smokers and ex- smokers not smoking due to flavours.

***Negative values*** indicate the number of smokers and ex-smokers who are not smoking due to flavours outweighs the number of youth smoking due to flavours.

Suggested Resources: ASH surveys (SmokeFree GB); England Smoking Toolkit Study; ONS Adult smoking habits in the UK (Annual Population Survey) E-cigarette use in England, NHS digital Smoking, Drinking and Drugs Survey (SDD); ONS Annual Population Survey (E-cigarette use in England) and Opinions and Lifestyle Survey (OPN)

### E-LIQUID POLICY DECISION AID

University of BRISTOL

#### OVERVIEW

This policy decision aid is designed to assist policymakers to make informed decisions regarding the restriction of flavoured e-liquids in the UK. This aid uses estimated population proportions to predict whether a total ban on e-liquid flavouring (in which only three e-liquids would remain on the market - unflavoured, tobacco and menthol) would result in the number of smokers in the population increasing or decreasing. It also estimates whether there would be more non-smoking youth who experiment with e-cigarette use as a result of flavoured e-liquid availability than smokers and ex-smokers who would not smoke due to flavoured e-liquid availability. The decision aid estimates a 'worst case scenario' whereby the negative impact on youth is likely overestimated and the negative impact on adult smokers and ex-smokers is likely underestimated. The quality of evidence for each estimated proportion entered into the decision aid may differ, which could impact the accuracy of the estimated impact. The notes section provides information about the quality of evidence entered into the aid.

#### IMPACT ASSESSMENT

The following impact assessment relates to UK General Population.

Using the available evidence on 20 February 2023, we estimate that 53,609 non-smoking youth experiment with e-cigarettes as a result of flavoured e-liquid availability and 328,403 smokers and ex-smokers do not smoke due to flavoured e-liquid availability. This output suggests that restricting flavoured e-liquids in the UK could have a negative overall impact on public health.

Using the available evidence on 20 February 2023, we estimate that 26,269 non-smoking youth subsequently smoke as a result of flavoured e-liquid availability and 328,403 smokers and ex-smokers do not smoke due to flavoured e-liquid availability. This output suggests that restricting flavoured e-liquids in the UK could have a negative overall impact on public health.

#### DISCLAIMER

The outputs resulting from the policy decision aid are only as good as the quality of evidence which is entered into the tool and existing evidence is limited. Much of the evidence is reliant on self-reported beliefs about how smokers and vapers think they will behave in various scenarios, but in the event of flavour restrictions being implemented, smokers' and vapers' actual behaviour may differ from their reported intentions. These estimates could therefore be an underestimate of the impact of e-liquid flavour restrictions on adult smokers and e-cigarette users. Also, while there is a strong association between e-cigarette use and later smoking, there is no evidence that e-cigarette use *causes* smoking among youth[1]. There is also evidence to suggest that at least part of this relationship could be explained by shared risk factors,[2] so it is likely that this decision aid will overestimate the number of youth who are at risk of smoking due to e-liquid flavour availability. Furthermore, the decision aid does not account for displacement (i.e., the number of youths who do not smoke because e-cigarettes are available)[3] [4].

The decision aid only assesses the direct impact on individuals' own smoking and vaping behaviour, not the impact on others (e.g., second-hand smoke or the effect of smoking role models). Although the impact on others is a concern, it is extremely difficult to quantify.

Where possible, we include UK- or GB-wide data and representative samples. However, some non-representative samples may be included, particularly when the estimates relate to high-risk populations. If non-representative samples are used, this will be made clear when the document is sent. Additionally, using advanced modelling methods could provide more accurate estimates. Data and advice resulting from the policy decision aid are applicable to the UK context and should not be used for policy decisions in other countries without consulting the designers of the policy decision aid.

#### HOW THE AID IS USED

Current evidence suggests that e-cigarettes are an effective smoking cessation tool for adults[5]. However, e-cigarette use is not limited to adult use (Figure 1). Some young people also use e-cigarettes, and there are concerns that e-cigarettes could act as a "gateway" to smoking among non-smoking youth (Figure 1). Although there is observational research which supports this possibility[6], there is limited evidence to suggest that this is currently happening in the UK.

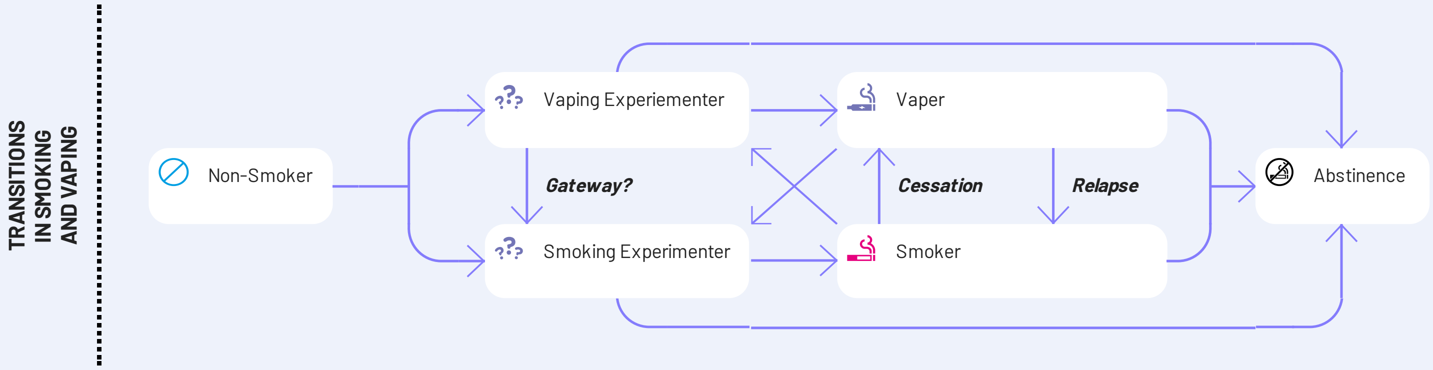

### E-LIQUID POLICY DECISION AID



Given the changing regulatory and product environment, it is important to continue monitoring the impact of e-cigarettes on youth and adult behaviour, in order to ensure that UK policy protects both youth and adults. To reduce youth uptake in some countries (e.g., the US), policymakers have attempted to make e-cigarettes less appealing to youth by restricting and banning flavours in e-liquids. These policies assume that flavours are attracting youth, and although some evidence does support this possibility, there is also evidence that some adult smokers and e-cigarette users are also attracted by the variety of flavour options. Therefore, restrictive policies could have unintended consequences such as fewer individuals using e-cigarettes to stop smoking or to prevent relapse.

This aid attempts to calculate the potential impact that restricting e-liquid flavours might have on the total number of smokers in the UK population, and the total number of youth e-cigarette users in the UK population.

The aid uses a basic algorithm in which upper case letters in the aid reflect:

A. The extent to which flavours draw non-smoking youth to vape.
B. How many non-smokers who vape as a result of e-liquid flavour availability may then go on to smoke.
C. To what extent e-liquid flavour availability draws in smokers who would quit smoking using e-cigarettes (per year).
D. The number of e-cigarette users who state they would relapse to smoking if flavoured e-liquid were not available on the UK market.

To estimate whether the number of non-smoking e-cigarette users introduced into the UK population because of flavoured e-liquid availability could outweigh the number of smokers and ex-smokers who might vape instead of smoke because of flavoured e-liquid availability, the following calculation is made:

$$A - (C + D)$$

To estimate whether the number of smokers introduced into the UK population because of flavoured e-liquid availability could outweigh the number of smokers and ex-smokers who might vape instead of smoke because of flavoured e-liquid availability, the following calculation is made:

$$B - (C + D)$$

Positive values would provide some support for a total ban on flavoured e-liquids, and negative values would provide some evidence that the negative consequences of a total ban would outweigh the potential benefits.

#### ALTERNATIVE POLICY OPTIONS

A total ban on flavoured e-liquids should be a last resort, particularly given the risks of creating a black market. Black market and unregulated products in the US appear to have caused over 60 deaths in the EVALI outbreak,[7] [8] and one in 10 e-cigarette users in the UK stated they would make their own flavours if they were to become unavailable.[9]

Restricting flavouring in e-liquids to products available only by prescription could reduce the impact of flavoured e-liquids on non-smoking youth while having a lesser impact on smokers and ex-smokers who would still be able to access these products via a prescription. However, independent e-cigarette companies must be supported to apply for medical licences to avoid a tobacco industry monopoly on flavoured e-liquids; as we have seen previously with the introduction of restrictions on e-cigarettes and nicotine concentrations in the UK, only the tobacco industry had sufficient funds to gain a medical licence, and they do not appear to be motivated to produce products for prescription. Medical professionals would also have to be encouraged to prescribe e-cigarettes as many are still cautious of e-cigarettes.

Flavour bans in other countries have omitted tobacco and menthol flavours. Any policy which restricts flavourings for use in e-cigarettes could consider restricting only the categories of flavour which appear to be most appealing to youth (e.g., alcoholic drinks, sweet/candy flavours).

Understanding the factors that may drive the appeal of flavoured e-liquids is also necessary to ensure the correct policy is implemented. If the flavour itself is the most attractive element of flavoured e-liquids, then a total ban would be advisable if the benefits outweigh the costs, however, flavoured products are also usually accompanied by attractive descriptors and appealing packaging which could be driving any effect of flavoured e-liquid use among youth. Restricting these factors may be sufficient to discourage youth use of the products while retaining adult users who would otherwise relapse to smoking and still attracting smokers who would stop smoking using e-cigarettes.

#### NOTES

#### CONTACT INFORMATION

University of BRISTOL

#### REFERENCES


1. Chan GCK, Stjepanović D, Lim C, et al. Gateway or common liability? A systematic review and meta-analysis of studies of adolescent e-cigarette use and future smoking initiation. Addiction 2020;n/a(n/a) doi: 10.1111/add.15246
2. Khouja JN, Wootton RE, Taylor AE, et al. Association of genetic liability to smoking initiation with e-cigarette use in young adults: A cohort study. PLoS Med 2021;18(3):e1003555. doi: 10.1371/journal.pmed.1003555 [published Online First: 2021/03/19]
3. Sokol NA, Feldman JM. High School Seniors Who Used E-Cigarettes May Have Otherwise Been Cigarette Smokers: Evidence From Monitoring the Future (United States, 2009-2018). Nicotine & Tobacco Research 2021;23(11):1958-61. doi: 10.1093/ntr/ntab102
4. Hallingberg B, Maynard OM, Bauld L, et al. Have e-cigarettes renormalised or displaced youth smoking? Results of a segmented regression analysis of repeated cross sectional survey data in England, Scotland and Wales. Tobacco Control 2020;29(2):207-16. doi: 10.1136/tobaccocontrol-2018-054584
5. Hartmann-Boyce J, McRobbie H, Lindson N, et al. Electronic cigarettes for smoking cessation. Cochrane Database of Systematic Reviews 2020(10) doi: 10.1002/14651858.CD010216.pub4
6. Khouja JN, Suddell SF, Peters SE, et al. Is e-cigarette use in non-smoking young adults associated with later smoking? A systematic review and meta-analysis. Tob Control 2020 doi: 10.1136/tobaccocontrol-2019-055433 [published Online First: 2020/03/12]
7. Werner AK, Koumans EH, Chatham-Stephens K, et al. Hospitalizations and Deaths Associated with EVALI. New England Journal of Medicine 2020;382(17):1589-98. doi: 10.1056/NEJMoa1915314
8. Ellington S, Salvatore PP, Ko J, et al. Update: Product, Substance-Use, and Demographic Characteristics of Hospitalized Patients in a Nationwide Outbreak of E-cigarette, or Vaping, Product Use-Associated Lung Injury — United States, August 2019-January 2020. MMWR Morb Mortal Wkly Rep 2020;69:44-49. doi: http://dx.doi.org/10.15585/mmwr.mm6902e2external
9. Action on Smoking and Health. Use of e-cigarettes (vapes) among adults in Great Britain, 2020.
